# Latent anxiety and depression dimensions differ among eating disorders: a Swedish nationwide investigation

**DOI:** 10.1101/2022.05.25.22275566

**Authors:** Christopher Hübel, Andreas Birgegård, Therese Johansson, Liselotte V. Petersen, Rasmus Isomaa, Moritz Herle

## Abstract

**Objective:** Anxiety and depression symptoms are common in eating disorders. To study these, we need high-quality self-report questionnaires. The 19-item self-rated Comprehensive Psychopathological Rating Scale for Affective Syndromes (CPRS-S-A) is not validated in eating disorders. We tested its factor structure, invariance, and differences its latent dimensions.

**Method:** Patients were registered by 45 treatment units in the Swedish nationwide Stepwise quality assurance database for specialised eating disorder care (*n* = 9,509). Patients self-reported their anxiety and depression symptoms on the CPRS-S-A. Analyses included exploratory and confirmatory factor analysis in a split samples and invariance difference testing in subscales across eating disorders.

**Results:** Results suggested a four-factor solution: Depression, Somatic & fear symptoms, Disinterest, and Worry. Multigroup confirmatory factor analysis indicated an invariant factor structure. We detected the following differences: Patients with anorexia nervosa binge-eating/purging subtype scored the highest and patients with unspecified feeding and eating disorders the lowest on all subscales. Patients with anorexia nervosa or purging disorder show more somatic & fear symptoms than individuals with either bulimia nervosa or binge-eating disorder.

**Conclusion:** Our four-factor solution of the CPRS-S-A is suitable for patients with eating disorders and may help to identify differences in anxiety and depression dimensions amongst eating disorders.

**Significant Outcomes:**

⍰ Symptoms of anxiety and depression in eating disorder patients load on four dimensions: Depression, Somatic & fear symptoms, Disinterest, and Worry.
⍰ Patients with anorexia nervosa binge-eating/purging subtype show the largest depression and anxiety symptom burden while patients with subsyndromal forms of eating disorders show the lowest.
⍰ Instead of calculating total scores of anxiety and depression, a dimensional approach delivers more fine-grained association results.

**Limitations:**

⍰ Our sample consisted of eating disorder patients only and there was no healthy control group or data from patients with other psychiatric disorders for comparison.
⍰ The sample consisted predominantly of women which limits the ability to identify sex differences.
⍰ The sample included Swedish treatment seeking patients of mostly white European ancestry limiting the generalisability of our findings.

## 1. Introduction

### 1.1 Comorbidity among affective disorders and eating disorders

Symptoms of anxiety and depression, such as disinterest, low mood, and suicidality are commonly found amongst patients with eating disorders in the population and clinic.^1–6^ Comorbid depression among individuals with any eating disorders is highly prevalent with 75%.^7^ Specifically, ∼60% of adolescents with anorexia nervosa report depressive symptoms^8,9^ and patients with anorexia nervosa often have a comorbid clinical depression.^10,11^ This high level of comorbidity with depression is also observed in individuals with bulimia nervosa, in population^12,13^ and clinical samples.^2,10,14^ Similarly to depression, anxiety and eating disorders also co-occur.^2,10,15–19^ However, prevalence estimates show considerable heterogeneity depending on measure or assessment type (e.g. self-report versus clinical interview).^20,21^ Overall, the co-occurrence of depressive or anxiety symptoms with eating disorder symptoms complicates treatment.^4,22,23^ Therefore, accurate assessment of anxiety and depression symptoms in eating disorders may be beneficial for treatment planning.

### 1.2 Measurement issues

To study comorbidity among eating disorders, anxiety, and depression, we need high-quality measures of symptoms to delineate differences in eating disorder presentation in clinical and population samples. Measures like the Patient Health Questionnaire-9 (PHQ-9)^24^ and the Generalised Anxiety Disorder Assessment^25^ are widely used in research. The PHQ-9 and GAD-7 factor structure and scores have been validated in samples of eating disorder patients and the general population, indicating that both questionnaires are suitable.^26^ However, as they are strictly based on diagnostic criteria, these questionnaires only cover a limited range of anxiety and depression symptoms. Hence, in order to explore the whole spectrum of symptoms, broader assessment tools to better understand the heterogeneity in eating disorder presentations, evading the cost- and time-related limitations of diagnostic interviews, are urgently needed. One potential scale of interest is the Comprehensive Psychopathological Rating Scale (CPRS)^27^ which consists of 65 items and was originally developed to evaluate treatment outcomes in psychological interventions. The scale includes items covering symptoms of psychiatric disorders, such as schizophrenia but also anxiety and depression. The scale was originally developed in Sweden, and has been translated into most other European languages. The complete version of the CPRS is rarely used, but shorter subscales have been deemed to be more useful, such as the Montgomery Åsberg Depression Rating Scale (MADRS)^28^ and the Self-rating Scale for Affective Syndromes (CPRS-S-A).^29^ The latter is in focus here, and is designed to contain subscales for depression, anxiety, and compulsivity.

A previous analysis of the CPRS-S-A questionnaire in a subsample of the data available for our investigation showed that patients with an unspecified feeding or eating disorder reported fewer problems than patients with other eating disorders. Additionally, patients with the anorexia nervosa binge-eating/purging subtype reported more problems compared with atypical anorexia nervosa patients.^30^ One issue of the questionnaire is the construction of its three subscales. When calculating the subscales, it is advised to include the same item in several subscales. Therefore, the subscales are highly correlated. In the previous analyses the correlations ranged from 0.78 to 0.86 and are inflated, rendering the original subscales unreliable. Therefore, in this study, we investigated differences in depression and anxiety dimension among eating disorders using newly derived subscales of the Self-rating Scale for Affective Syndromes (CPRS-S-A); a short form of the CPRS^29^ in one of the World’s largest clinical samples of more than 9,000 individuals with eating disorders in Sweden.

## 2. Methods

### 2.1 Sample

The sample comprises inpatients and outpatients registered by 45 treatment units in the Stepwise quality assurance database for specialised eating disorder care in Sweden aged 18 years and older.^31^ Stepwise is a nationwide internet-based data collection system, which includes individuals through medical or self-referral, if intention to treat has been established, and if the individual received a formal eating disorder diagnosis.^32^ The database has been used since 2005 and our data were extracted on 23^rd^ of November 2017. At data extraction, approximately 10,470 adult patients had been registered.

### 2.2 Eating disorder diagnosis

Clinicians registered patients’ eating disorders diagnosis based on Diagnostic and Statistical Manual of Mental Disorders, Fourth Edition (DSM-IV).^32,33^ In our analysis, we translated DSM-IV to DSM-5 eating disorders to reflect the current understanding of eating disorders. Depending on the patient’s endorsement of binge eating or purging in either the Eating Disorder Examination questionnaire (EDE-Q)^34^ or the Structured Eating Disorder Interview (SEDI),^35^ we re-assigned DSM-5 diagnoses. We used 18.5 kg/m^2^ as the cutoff value for underweight in anorexia nervosa. Anorexia nervosa without weight criterion (*n* = 50) or without amenorrhea (*n* = 186) that had a BMI lower than 18.5 kg/m^2^ who endorsed any binge eating or purging were assigned anorexia nervosa binge-eating/purging. If none endorsed, they were assigned an anorexia nervosa restricting diagnosis (*n*_without weight criterion_ = 84 or *n*_without amenorrhea_ = 144). If their BMI was above 18.5 kg/m^2^, we assigned an atypical anorexia nervosa diagnosis (*n*_without weight criterion_= 441 or *n*_without amenorrhea_ = 402). Eating Disorder Not Otherwise Specified (EDNOS) type 3 or bulimia nervosa without sufficient duration/frequency criteria (*n* = 833) was assigned as bulimia nervosa diagnosis because those criteria are relaxed in DSM-5. Further, EDNOS example 4 was kept as “purging disorder”. The remaining unspecified eating disorders that were not classified into either of these categories were termed “Unspecified feeding or eating disorder” (UFED), consisting of patients with “chewing and spitting”, bulimia nervosa/binge-eating disorder with low frequency/duration, or other residual types that did not fit any of the main categories.

### 2.3 Exclusion

We excluded 801 duplicated entries of repeated registrations of the same individual. Subsequently, we iteratively excluded 2 individuals with missing age, 16 not assigned a treatment centre, 120 without a clinical eating disorder diagnosis, and 22 because they had not answered the CPRS questionnaire. The final sample comprised 9,509 eating disorder patients.

### 2.4 Ethics

When patients were entered into the database, clinicians recorded consent for general research use of their data and 3% declined participation. This study is approved by the Stockholm Regional Ethics Board (Reg. no. 2009/196-31/4).

### 2.5 Comprehensive Psychopathological Rating Scale, Self-rated Version for Affective Syndromes

At registration, the patients answered 19 items of the CPRS-S-A. We present the instrument as **Supplementary Material**. The answer options are different for each question, but they are on a scale from 0 to 3, rated in 0.5-point increments. We recoded these values to 0–6. We renamed Iiem 19, titled “Zest for life” in the MADRS-S to “Suicidal thoughts” to represent its content better.

### 2.6 Exploratory factor analyses

We calculated pairwise Pearson correlations among all items (**Figure 1**) in the full sample (*n* = 9,509). We inspected the matrix visually for singularity, multicollinearity, and redundancy of items (i.e., values <0.30 and >0.90). We calculated the determinant of the matrix,^36^ the Kaiser-Meyer-Olkin (KMO) statistic,^37^ and performed Bartlett’s Test of Sphericity,^38^ to test if our data are suitable for an exploratory factor analysis. To inform our decision on the underlying factor structure, we performed parallel analysis,^39^ and calculated the Very Simple Structure criterion (VSS),^40^ and Velicer’s Minimum Average Partial (MAP) criterion.^41^ We performed the exploratory factor analysis on 70% (*n* = 6,656) of the sample using the maximum likelihood estimator in the “psych” R package.^42^ Given that the CPRS items have seven answer options, we treated them as continuous. We allowed the factors to correlate using oblimin rotation. To judge the fit of our model, we applied the criteria as outlined in **Table 1**.^43^ We retained the items with factor loadings of >0.30. If multiple models showed adequate fit, we would choose the model with factors that encompass the greatest number of items.

**Figure 1.**
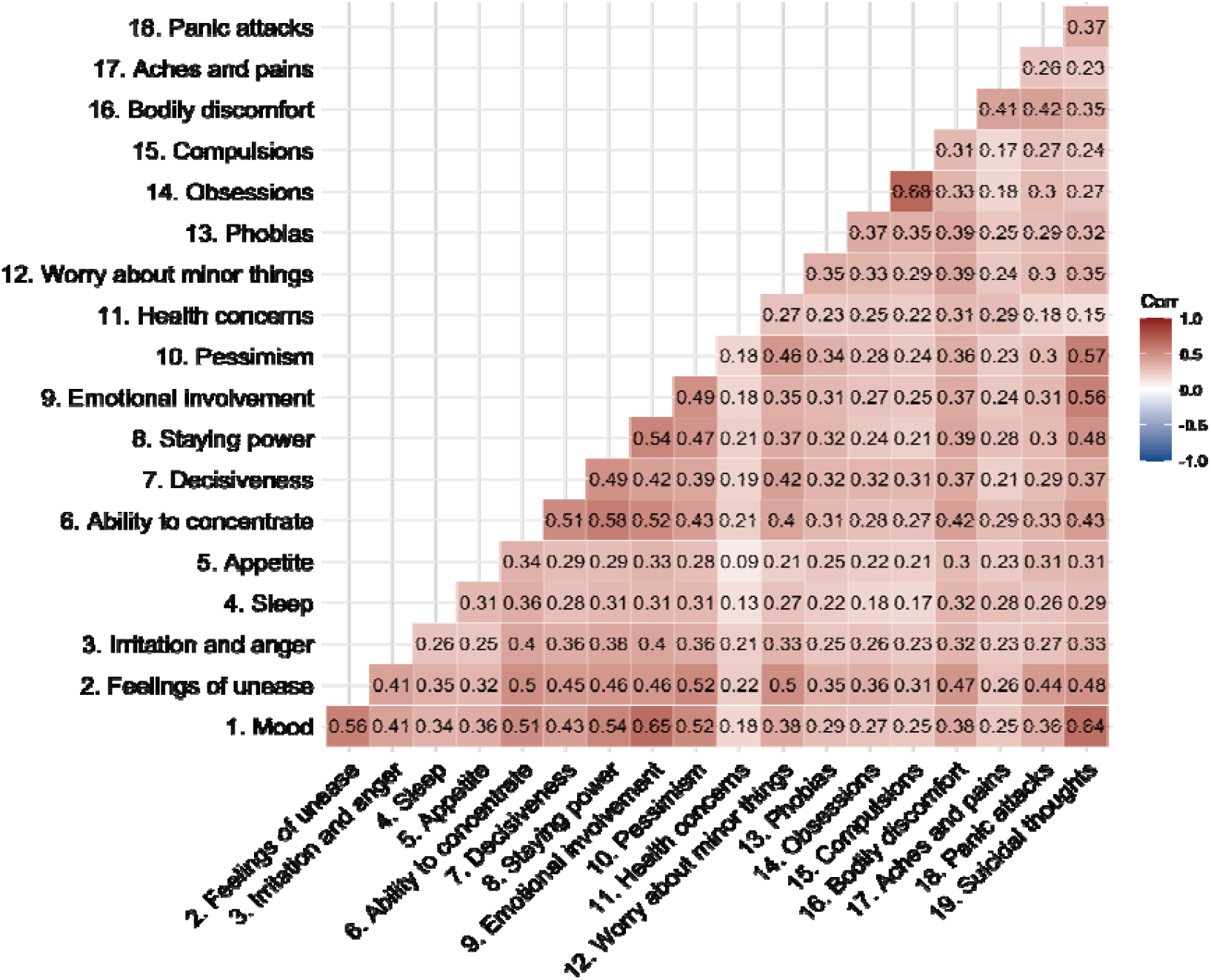
Pairwise Pearson’s correlations among the Self-rating Scale for Affective Syndromes (CPRS-S-A) items. We calculated the correlations in 9,509 participants registered in Stepwise, the Swedish clinical eating disorder database. We estimated the number of independent traits in the matrix using the Galwey method and adjusted the ⍰ threshold (⍰ = 0.003) accordingly. All correlations are statistically significant at this ⍰ threshold. Saturation represents the strength of the correlation. Positive correlations are red, negative correlations are blue.

**Table 1.** Criteria for a good fit^43^

|  |  |
| --- | --- |
| Root Mean Square Error of Approximation (RMSEA) | $\leq 0.05$ |
| Tucker Lewis Index (TLI) | $\geq 0.95$ |
| Standardised Root Mean Square Residuals (SRMR) | $\leq 0.05$ |
| Bayesian Information Criteria (BIC) | Smaller than other models |

### 2.7 Confirmatory factor analysis and factor scores

We validated our exploratory factor analysis model with a confirmatory factor analysis (CFA) on the remaining 30% participants using the “lavaan” R package.^44^ We interpreted fit statistics^43,45^ and considered a Comparative Fit Index (CFI) ≥0.95 as good fit. Subsequently, we computed the confirmatory factor analysis in the full sample (*n* = 9,509) to provide fit statistics and calculate factor scores, using the Bartlett estimator for continuous items.

### 2.8 Descriptive indices and psychometric properties

We show responses to the individual items and distributions of the factor scores as frequency and box plots. We also report mean and standard deviations for our generated factor scores and report Cronbach’s ⍰^46,47^ and McDonald’s ⍰^48^ as measures of internal consistency.

### 2.9 Multigroup confirmatory factor analysis

We performed a multigroup confirmatory factor analysis (MGCFA) to test if the questionnaire elicits the same responses, response patterns, and has the same underlying factor structure across eating disorder diagnostic groups. If statistical invariance in responding is found, then we can compare scores and subscale scores across groups. Different types of measurement invariance exist: configural, the factor structure is similar across groups; metric, factor loadings are similar across groups; scalar, intercepts (i.e., group means) are similar; and strict, residuals (i.e., variances) are similar across the groups. We tested for these invariance models in a stepwise procedure from the least restricted model to the fully restricted model. Overall, invariance indicates that different groups are from the same population.

### 2.10 Group comparisons

We judged the distribution of the factor scores by visually inspecting qq and distribution plots (**Supplementary Figure S1**). None of the four subscales showed a normal distribution. Therefore, we performed non-parametric Kruskal-Wallis one-way ANOVAs. If significant, Dunn’s post-hoc tests were carried out with a Benjamini Hochberg-adjusted level of significance for the pairwise comparisons.

### 2.11 Convergent and divergent validity

If our newly developed dimensions correlate with other instruments that measure similar constructs in the expected direction and with sufficient magnitude, they show convergent validity. To assess this, we estimated correlations with the original depression and anxiety subscales of the CPRS-S-A, the Clinical Impairment Assessment (CIA) total score^49^ (both expected to be positive), and the Structural Analysis of Social Behavior (SASB) self-affirmation scale (expected to be negative).50 For divergent validity, we correlated the new CPRS-S-A dimensions with variables that we expected to be unrelated: SASB self-control and height.

## 3. Results

### 3.1 Descriptives

The patients in our sample were on average 26 years (*SD* = 8) old and the age ranged from 18 to 70 years, with 96% of the sample being female. Of the patients, 1,363 (14%) received an anorexia nervosa restricting, 702 (7%) anorexia nervosa binge-eating/purging, 832 (9%) an atypical anorexia nervosa, 3,807 (40%) a bulimia nervosa, 658 (7%) a binge-eating disorder, 1,711 (18%) a purging disorder, and 436 UFED diagnosis (5%).

### 3.2. Descriptives of the CPRS-S-A in Stepwise

The CPRS-S-A showed a Cronbach’s ⍰ (⍰ = 0.90) and McDonald’s ⍰ (⍰ = 0.92) in our sample (**Supplementary Table S1**). The distribution of answers to the questionnaire items is displayed in **Figure 2** for the full sample and **Supplementary Table S2** for the discovery sample.

**Figure 2.**
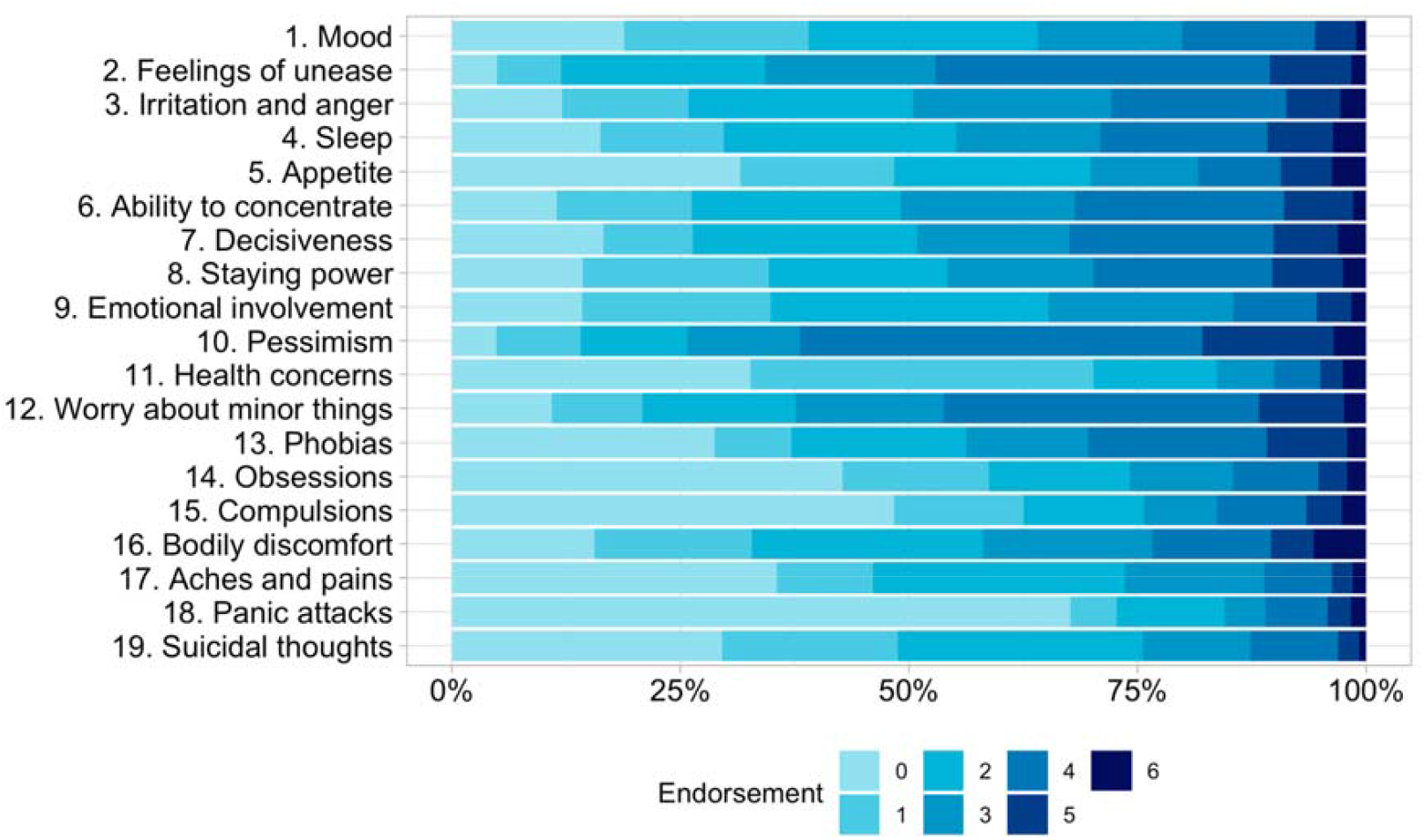
Endorsement of the Comprehensive Psychopathological Rating Scale Self-rating Scale for Affective Syndromes (CPRS-S-A) 19 items version in the Stepwise sample (n = 9,509). The saturation of blue indicates a higher endorsement on the specific item. We display percentages. Originally, each item had seven answer options ranging from 0-3, which were later recorded from 0-6 for the main analyses. The answer options differed across items, with higher values indicating a stronger endorsement.

### 3.3 Suitability of the data for factor analysis

Prior to factor analyses, the suitability of the data was investigated. None of the items showed zero or near-zero variance (**Supplementary Table S3**). Kaiser-Meyer-Olkin measure (KMO = 0.94, **Supplementary Table S4**) and significant Bartlett test of sphericity (*p* < 2.22 × 10^−16^) indicated that the data were suitable for factor analyses. Pearson’s correlations ranged from 0.09 to 0.68 (Figure 1). The exploratory factor analysis was conducted on one random split of the sample (*n* = 6,656; 70%). As we were primarily interested in core anxiety and depression symptoms, we excluded the items “14. Obsessions” and “15. Compulsions” from the factor analysis. Furthermore, they loaded strongly on one factor by themselves, representing an index of compulsion. If these items had remained in the model, they would have lowered our power to measure meaningful underlying factors as they would have distorted the model towards their own factor. We, furthermore, excluded the item “11. Health concerns”, because its correlation with the other items was small (*r* = 0.09 to 0.29; **Figure 1**), rendering it unsuitable for factor analysis. Cronbach’s ⍰ remained stable after these items were dropped (**Supplementary Table S5**)

### 3.4 Exploratory factor analysis

Very simple structure (**Supplementary Table S6**) and parallel analysis (**Supplementary Table S7**) suggested a one factor solution. However, as we are interested in different anxiety and depression symptoms, a comparison of fit statistics suggested that the five factor solution fitted the data best. However, the model contained two factors on which only one item (i.e., 2. Feelings of unease & 12. Worry about minor things) and therefore the model was unsuitable. Hence, we chose the four-factor solution as our final model which explained 34% of the total variance. The factor solution had a low RMSEA (0.042, 90% CI: 0.039 0.045) and low Bayesian Information Criterion (BIC = 245; for full results, see **Table 2 and Supplementary Tables S8-12**). As factors were considered to be correlated, factors were realigned using an oblique rotation. The factor loadings for each item, after rotation, are listed in **Figure 3**. Items 3 and 4 (Irritation and anger, and Sleep, respectively) did not load on any of the factors and are therefore not included in the confirmatory factor analysis. We labelled the four factors: F1 Depression, F2 Somatic & fear symptoms, F3 Disinterest, and F4 Worry.

**Table 2.** Model fit statistics for exploratory factor analysis. The factor analysis was performed on 16 items of the Comprehensive Psychopathological Rating Scale Self-rating Scale for Affective Syndromes (CPRS-S-A) in the Swedish quality register for eating disorder care, Stepwise (n = 6,656).

| Number of factors | df | RMSEA ($\leq 0.06$) | RMSEA 90% CI | TLI ($\geq 0.95$) | BIC | SRMR ($\leq 0.08$) | Cumulative variance | Minimum item loading |
| --- | --- | --- | --- | --- | --- | --- | --- | --- |
| 1 | 104 | 0.078 | [0.076, 0.080] | 0.882 | 3351 | 0.05 | 0.38 | 16 |
| 2 | 89 | 0.062 | [0.060, 0.065] | 0.923 | 1608 | 0.03 | 0.35 | 7 |
| 3 | 75 | 0.053 | [0.051 0.056] | 0.944 | 840 | 0.03 | 0.33 | 3 |
| **4** | **62** | **0.042** | **[0.039 0.045]** | **0.965** | **245** | **0.02** | **0.34** | **2** |
| 5 | 50 | 0.035 | [0.033 0.038] | 0.975 | 29 | 0.01 | 0.34 | 1 |

df: Degrees of freedom; RMSEA: Root Mean Square Error of Approximation; TLI: Tucker-Lewis Fit Index; BIC: Bayesian Information Criterion; SRMR: Standardised Root Mean Square Residuals. The cut off for each statistic to signify good fit is listed in each header (Hu & Bentler, 1999). The model with the lowest BIC is preferred. Cumulative variance indicates the part of the total variance explained by all items comprising the factors.

**Figure 3.**
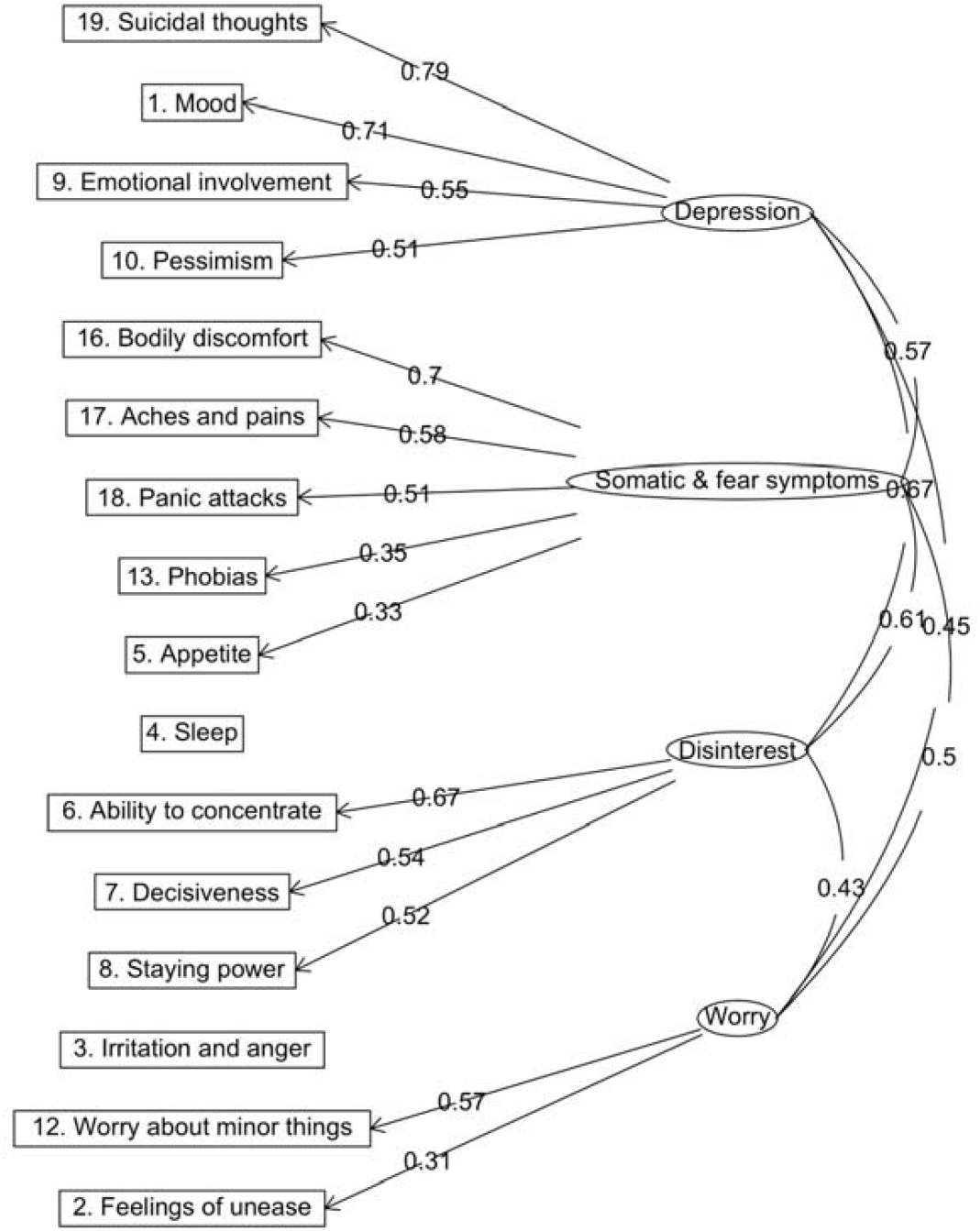
Exploratory factor analysis of 16 items of the Comprehensive Psychopathological Rating Scale Self-rating Scale for Affective Syndromes (CPRS-S-A). The path diagram shows item factor loadings and between-factor correlations for the four factors of Depression, Somatic & fear symptoms, Disinterest, and Worry. Paths with a factor loading of <0.3 were omitted.

### 3.5 Confirmatory factor analysis

We conducted the confirmatory factor analysis in the remaining 30% of the sample (*n* = 2,853; **Supplementary Table S13**). Results confirmed that the four-factor model was a good fit. The RMSEA (0.060, 90% CI: 0.056, 0.064), the CFI (0.952), and the SRMR (0.032) indicated good fit.^43^ The TLI (0.939) was slightly above the threshold for good fit. We also ran the confirmatory factor analysis in the full sample (*n* = 9,509) which yielded the following fit statistics: CFI = 0.953, TLI = 0.940, RMSEA = 0.060 [90% CI, 0.058, 0.062], SRMR = 0.030). We show the resulting factor scores and their distribution in Figure 4 and Supplementary Figure S1.

**Figure 4.**
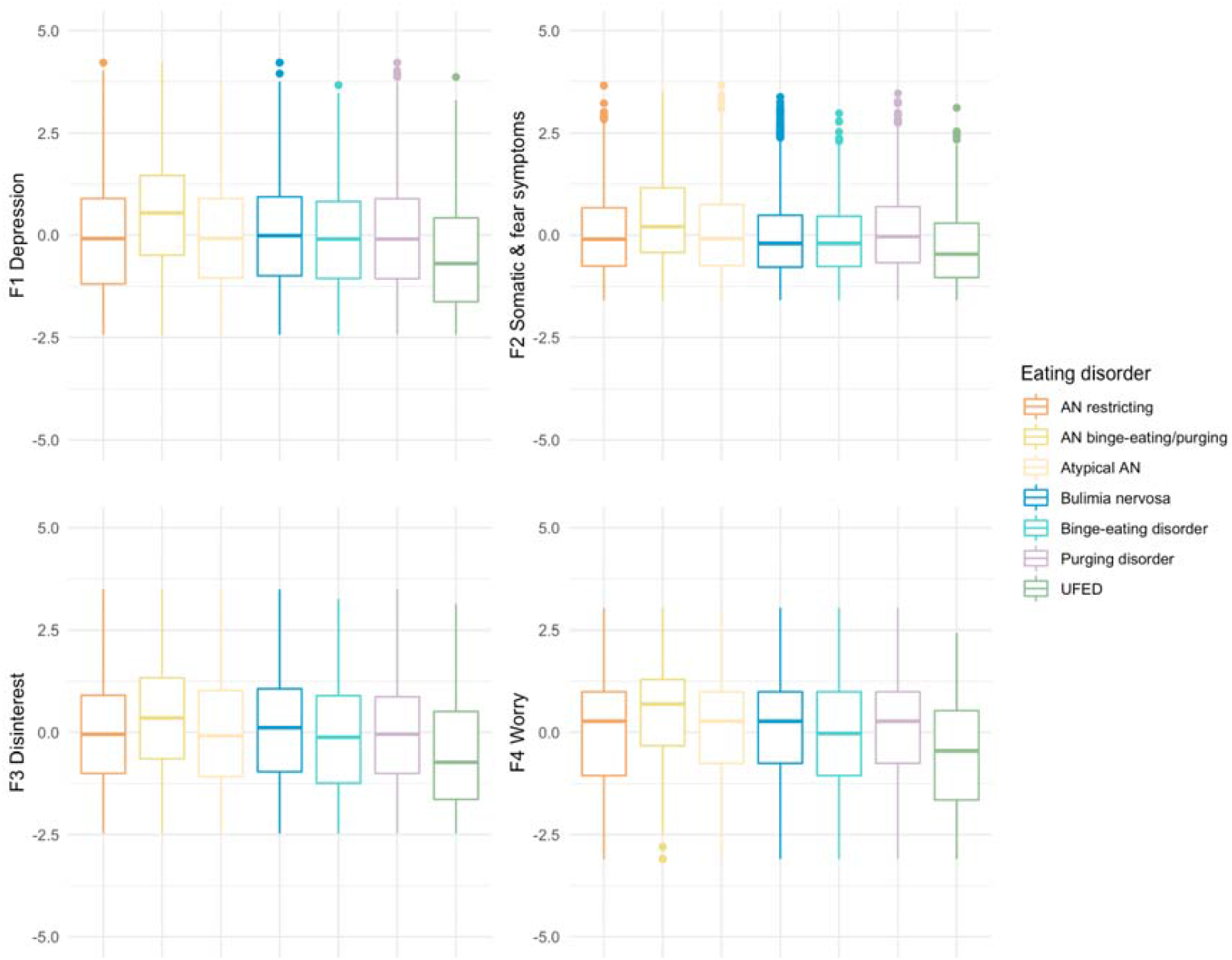
Distribution of factor scores across eating disorders. Boxplots represent median and interquartile range in the whole Stepwise sample (n = 9,509). Note. AN, anorexia nervosa, UFED, unspecified feeding or eating disorder

### 3.6 Multigroup confirmatory factor analysis

Our multigroup confirmatory factor analysis resulted in full configural and metric invariance, indicating that the factor structure and the factor loadings are comparable across eating disorders (**Supplementary Table S14**). Furthermore, the questionnaire showed partial scalar invariance when freeing up the intercepts of item five and eight, meaning that the means were similar across groups apart from item five (less appetite) and eight (less motivation).

### 3.7 Factor scores

We calculated factor scores for each individual based on the final model (**Figure 4 & Supplementary Table S15**). We compared the factor scores using Kruskal–Wallis one-way analysis of variance, and Dunn’s post hoc test (**Table 3**). Overall, individuals with anorexia nervosa binge-eating/purging scored higher on all four subscales than all other eating disorders, including the restricting subtype of anorexia nervosa. However, there was no statistically significant difference on any of the four scores between the restricting subtype and atypical anorexia nervosa or purging disorder. Individuals with UFED scored lower on all four scales than all other eating disorders.

**Table 3.** Median differences and results from Dunn’s post-hoc tests. We performed Kruskal-Wallis one-way Analysis of Variance (ANOVA) and Dunn’s post-hoc tests for pairwise comparisons and judged significance by adjusting the alpha level using the Benjamini Hochberg approach.

| Factor | ANR<br>vs.<br>ANBP | ANR<br>vs.<br>AAN | ANR<br>vs.<br>BN | ANR<br>vs.<br>BED | ANR<br>vs.<br>PUR | ANR<br>vs.<br>UFED | ANBP<br>vs.<br>AAN | ANBP<br>vs.<br>BN | ANBP<br>vs.<br>BED | ANBP<br>vs.<br>PUR | ANBP<br>vs.<br>UFED | AAN<br>vs.<br>BN | AAN<br>vs.<br>BED | AAN<br>vs.<br>PUR | AAN<br>vs.<br>UFED | BN<br>vs.<br>BED | BN<br>vs.<br>PUR | BN<br>vs.<br>UFED | BED<br>vs.<br>PUR | BED<br>vs.<br>UFED | PUR<br>vs.<br>UFED |
| --- | --- | --- | --- | --- | --- | --- | --- | --- | --- | --- | --- | --- | --- | --- | --- | --- | --- | --- | --- | --- | --- |
| F1 | 0.6**** | 0.0 | 0.1 | 0.0 | 0.0 | -0.6**** | -0.6**** | -0.5**** | -0.6**** | -0.6**** | -1.2**** | 0.1 | 0.0 | 0.0 | -0.6**** | -0.1 | -0.1 | -0.7**** | 0.0 | -0.6**** | -0.6**** |
| F2 | 0.3**** | 0.0 | -0.1** | -0.1 | 0.1 | -0.4**** | -0.3**** | -0.4**** | -0.4**** | -0.2**** | -0.7**** | -0.1*** | -0.1** | -0.1** | -0.4**** | 0.0 | 0.2**** | -0.3**** | 0.2** | -0.3*** | -0.5**** |
| F3 | 0.3**** | -0.1 | 0.1* | -0.1 | 0.0 | -0.7**** | -0.4**** | -0.2**** | -0.4**** | -0.3**** | -1.0**** | 0.2 | 0.0 | 0.0 | -0.6**** | -0.2** | -0.1** | -0.8**** | 0.1 | -0.6**** | -0.7**** |
| F4 | 0.4**** | 0.0 | 0.0 | -0.3* | 0.0 | -0.8**** | -0.4**** | -0.4**** | -0.7**** | -0.4**** | -1.2**** | 0.0 | -0.3* | -0.3* | -0.8**** | -0.3** | 0.0 | -0.8**** | 0.3** | -0.5**** | -0.8**** |

**Note.** F1 Depression, F2 Somatic & fear symptoms, F3 Disinterest, and F4 Worry, \*\*\*\* < $1 \times 10^{-4}$ , \*\*\* < 0.001, \*\* < 0.01, \* < 0.05
ANR, anorexia nervosa restricting; ANBP, anorexia nervosa binge-eating/purging; AAN, atypical anorexia nervosa, BN, bulimia nervosa; BED, binge-eating disorder, PUR, purging disorder patients, UFED, unspecified feeding or eating disorder.

On factor 2 Somatic & fear symptoms, patients with anorexia nervosa or purging disorder scored higher than individuals with either bulimia nervosa or binge-eating disorder. Patients with anorexia nervosa restricting subtype or atypical anorexia nervosa reported depressive symptoms on the same median level as patients with either bulimia nervosa, binge-eating disorder, or purging disorder. However, compared with anorexia nervosa binge-eating/purging, all other eating disorders reported fewer depressive symptoms. On factor 3 Disinterest, the results showed a mixed picture: patients with either anorexia nervosa binge-eating/purging subtype or bulimia nervosa reported more disinterest than the other eating disorders.

### 3.8 Convergent and divergent validity

Correlations between the original CPRS subscales and the new factors were positive and high (range r = 0.65 to 0.91), indicating that they measure similar underlying constructs (**Figure 5**). Importantly, the correlations between the new dimensions (range *r* = 0.51 to 0.68) were lower than between the original CPRS scales of depression and anxiety (*r* = 0.78), indicating that the new dimensions measure diverging underlying constructs. As hypothesised, the new CPRS dimensions were not correlated with either SASB self-control (range *r* = -0.02 to 0.08) or height (range *r* = -0.05 to -0.02), but were positively correlated with the CIA total score (range *r* = 0.51 to 0.61) and negatively with self-affirmation (range *r* = -0.54 to -0.34)

**Figure 5.**
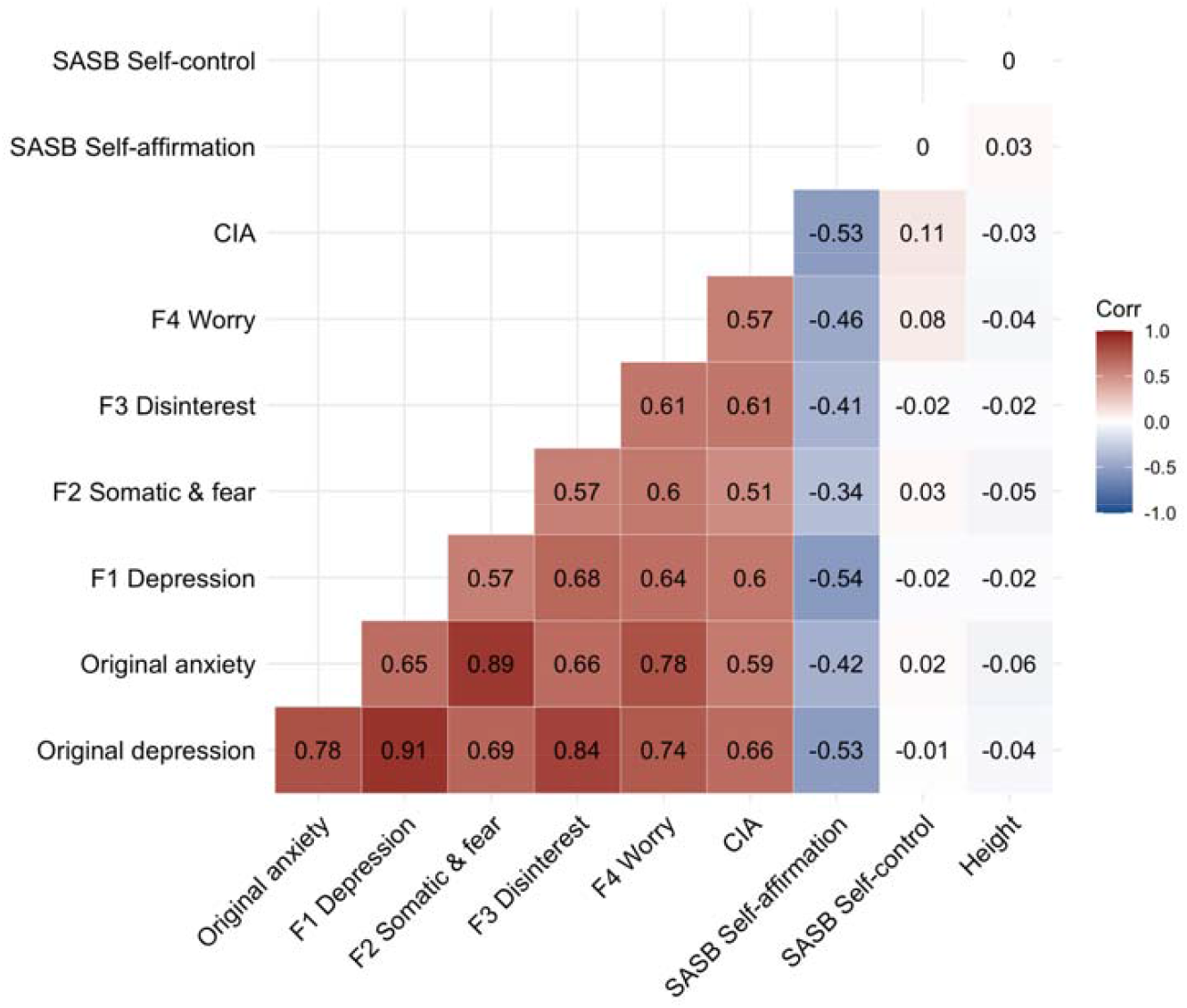
Correlation matrix of CPRS-S-A dimensions with external correlates. The correlation matrix shows Pearson’s correlations of the original CPRS-S-A anxiety and depression dimensions, the newly derived 4 factor solution, the Clinical Impairment Assessment (CIA) total score, and both subscales of the Structural Analysis of Social Behavior (SASB) self-affirmation and self-control als well as height. Sample sizes range between 8,146 and 9,509.

## 4. Discussion

### 4.1 Summary

#### Summary of findings

Using a factor analytic approach, we propose and confirm a four factor structure of the Self-rating Scale for Affective Syndromes (CPRS-S-A) in the world’s largest clinical sample of 9,509 patients with eating disorders. The four dimensions capture specific aspects of depression and anxiety: 1. Depression (4 items), 2. Somatic & fear symptoms (5 items), 3. Disinterest (3 items) and 4. Worry (2 items). Furthermore, we show that the new subscales can be used to measure differences in these dimensions among eating disorders.

#### Reduced scale

Our psychometric analysis suggests items can be removed due to their low correlations with other items. Our proposed reduced scale has a total of 14 questionnaire items and is hence shorter than the original CPRS-S-A with 19 items. First, we could drop item “11. Health concerns” as it barely correlated with other items on the scale. Second, we dropped two items regarding compulsiveness (i.e., item 14 & 15) as these were not deemed core to depression and anxiety, were highly correlated with each other.. Further, two items (i.e., 4. Sleep and 3. Irritation and anger) did not sufficiently load onto any of our four factors and were hence dropped.

### 4.2 Context of existing literature

#### Difference to original scale

Our factor structure differs substantially from the original CPRS-S-A ^29^. In contrast to the original structure (**Supplementary Table S16**), our new structure splits traditional depression symptoms into separate dimensions: depression and disinterest. Depression mostly included indicator items of low mood, pessimism, and lack of enjoyment, whereas Disinterest revolved around cognition, such as lack of concentration and decision making. Anxiety symptoms were also split into two factors. First, the Somatic & fear symptoms factor grouped together general pain, bodily discomfort, physical panic attack symptoms, such as heart palpitations and dizziness, with phobias which can present with somatic symptoms. The factor also included an item probing differences in appetite. This item may be inappropriate in the context of eating disorders as changes in appetite can be a central symptom of eating disorders; however, the changes may be diametral depending on the eating disorder type. The appetite item loaded poorly on its factor and may be removed at the researcher’s or clinician’s discretion. Second, the Worry factor groups General worry about minor things and Feeling of unease together. This differs from the original CPRS-S-A that combined worry symptoms with the somatic and fear-based symptoms. Overall, our analyses suggest a substantially different factor structure compared with the original structure.

#### General differences amongst eating disorders

We explored differences in the new subscales amongst eating disorders. On the one hand, comparisons suggest that patients with anorexia nervosa binge-eating/purging score higher on all four subscales, consistent with the previous report based on a subsample of our analysis,^10^ on the other hand, unspecified feeding and eating disorder patients had the lowest scores across all four subscales in line with their subsyndromal expression of eating disorders.

#### Specific differences

In addition to these overarching differences, we detected differences for specific factors. On factor 2 Somatic & fear symptoms, patients with anorexia nervosa or purging disorder scored higher than individuals with either bulimia nervosa or binge-eating disorder. These differences may indicate that the somatic complications seen in anorexia nervosa^51^ and purging disorder may be captured by items on this factor summarising somatic fear symptoms. Furthermore, patients with anorexia nervosa and purging disorders may perceive these somatic and fear symptoms more strongly than patients with bulimia nervosa or binge-eating disorder. Fear has been proposed as a fundamental mechanism in the development of anorexia nervosa.^52^ Depression and anxiety are risk factors for eating disorders,^19,21^ but certain symptoms of anxiety or depression may represent somatic or psychiatric complications or sequelae of the eating disorder itself. However, in some cases, depressive and anxiety symptoms may be independent of the eating disorder. This underscores the importance of investigating anxiety and depression on the dimension or symptom level rather than using total scores.

### 4.3 Limitations

Our study may be biassed due to limitations. The sample consisted predominantly of women which limits the ability to identify sex differences. Eating disorders are more commonly diagnosed among women, however, men are underrepresented in eating disorder research. This may be due to a lack of awareness and understanding for these disorders among the wider community and clinicians or may represent an underlying true sex difference. Our sample included Swedish treatment seeking patients of mostly white European ancestry limiting the generalisability of our findings. Furthermore, patients in healthcare registers may represent a more severe subpopulation of individuals with eating disorders. Hence, the factor structure and our observed differences amongst eating disorders may not replicate across other ancestry or cultural groups or in individuals with less severe presentations. Our analyses were cross-sectional.

### 4.4 Future directions

To address a few of our limitations, future studies should confirm our newly detected factor structure in community samples, samples with other psychiatric disorders, and include a healthy comparison group. Optimally, researchers would collect repeated measures of the CPRS-S-A that would further our understanding of how these constructs develop over time and how levels of depression, disinterest, fear, and worry may change with treatment. Future studies could investigate clinical cutoffs to measure comorbid depressive and anxiety disorders.

### 4.5 Conclusions

In summary, our four factor solution of the CPRS-S-A is suitable for adult patients with different eating disorders and identifies differences in anxiety and depression dimensions. An easily administered, reliable self-report measure for the most common forms of comorbidity in eating disorders is clinically and for research important. The CPRS-S-A may aid the clinician in case formulation and treatment planning. It may also be relevant for the patient’s own understanding of their situation. A discussion between patient and clinician, facilitated by the individual CPRS-S-A results, of depression and anxiety dimensions or symptoms in relation to eating disorder symptoms may improve therapeutic alliance and thus treatment outcome.

## Data Availability

Due to regional legal regulations, the data from the quality register cannot be shared.

## Code availability

Code will be available on https://github.com/topherhuebel/CPRS-SA-ED.

## Conflict of Interest Statement

The authors report no conflict of interest.

## Acknowledgement

Liselotte V. Petersen and Christopher Hübel acknowledge funding by Lundbeckfonden (R276-2018-4581). Moritz Herle is funded by a fellowship from the Medical Research Council UK (MR/T027843/1).

## Authors contribution statement

Christopher Hübel: Conceptualisation, Software, Visualisation, Formal analyses, Writing - Original Draft, Writing - Review & Editing,. Andreas Birgegård: Data Curation, Supervision, Writing - Review & Editing, Funding acquisition, Resources, Therese Johansson: Software, Writing - Review & Editing. Rasmus Isomaa: Supervision, Writing - Review & Editing, Liselotte V. Petersen: Supervision, Writing - Review & Editing, Funding acquisition, Moritz Herle: Supervision, Conceptualisation, Writing - Original Draft, Writing - Review & Editing, Project administration

## Notes

### Competing Interest Statement

The authors have declared no competing interest.

### Summary of Updates

We added external validiation to the manuscript.

