## Supplementary Material for "Latent anxiety and depression dimensions differ among eating disorders: a Swedish nationwide investigation"

**Supplementary Methods**

**Clinical impairment assessment (CIA)**

The 16-item questionnaire measures the severity of secondary psychosocial impairment due to eating disorder features during the previous 28 days, covering three domains: emotional, social, and cognitive functioning. Each item is rated on a 4-point Likert scale with 0 = ‘not at all’, 1 = ‘a little’, 2 = ‘quite a bit’, and 3 = ‘a lot’. Summing all items yields a total severity score ranging from 0 to 48, with higher scores indicating greater impairment (Bohn et al., 2008).

**Structural analysis of social behavior (SASB)**

Structural analysis of social behavior (SASB) is a model that can be used to assess interpersonal and intrapsychic interactions in terms of three underlying dimensions: (a) focus (other, self, introject), (b) affiliation-hostility (love-hate), and (c) interdependence-independence (enmeshment-differentiation). Assessment of individuals or groups in terms of these dimensions can be made by self-ratings on the SASB Intrex questionnaires (Benjamin, 1974). We used the self-affirmation and the self-control subscales of the questionnaire.

**Supplementary Table 16. A comparison of the original factors and the newly derived factors.**

The table summarises the items comparing the original scale to the newly derived subscales from our exploratory factor analysis

Original scale: A: anxiety, D: depression, C: compulsion

Excluded: C: compulsion index

F1: Depression (D), F2: Somatic & fear symptoms (SF), F3: Disinterest (Di), F4: Worry (W)

| **Original scale** | **EFA** | **Item** |
| --- | --- | --- |
| D, C | D | **1. Mood.** Here you should try to indicate your mood, whether you have felt sad or gloomy. Try to recall how you have felt during the past 3 days, whether your mood has been changeable or much the same all the time. In particular, try to recall whether you have felt more cheerful if something good happened. |
| D, A, C | W | **2. Feelings of unease.** Here you should indicate to what extent you have had feelings of inner tension, uneasiness, anxiety, or vague fear, during the past 3 days. Pay particular attention to how intense any such feelings have been, whether they have come and gone or persisted almost all the time. |
| A |  | **3. Irritation and anger.** The question concerns how irritated or angry you feel inside, regardless of whether you have shown it or not. Think especially of how easily your anger flares up in relation to what triggered it, and how often and how intensely you felt angry or irritated. If you don’t have any such feelings at all, mark zero. |
| D, A |  | **4. Sleep.** Here you should indicate how well you sleep – how long you sleep, and how good your sleep has been for the past 3 nights. Your assessment should reflect how you have actually slept, regardless of whether you have used sleeping pills. If you have slept more than usual, you should mark the scale at zero. |
| D | SF | **5. Appetite.** Here you should indicate how your appetite has been, and try to recall whether it has differed in any way from normal. If your appetite has been better than usual, you should mark the scale at zero. |
| D, C | Di | **6. Ability to concentrate.** Here you should try to indicate your ability to collect your thoughts, to concentrate on what you are doing. Try to recall how well you have been able to cope with tasks requiring different degrees of concentration – for instance, compare your ability to read a more complex text and an easy passage in the newspaper, or to pay attention to the TV. |
| C | Di | **7. Decisiveness.** The question concerns your ability to make decisions in common, simple everyday situations, such as answering uncomplicated questions, or choosing between alternatives, for example deciding what to wear, what to have for dinner, or which TV show to watch. |
| D, C | Di | **8. Staying power.** Here you should try to assess your staying power, and whether you feel you tire easier than usual. |
| D | D | **9. Emotional involvement.** Here you should assess your interest in your surroundings, in other people, and in activities that normally give you pleasure. |
| D | D | **10. Pessimism.** Here you should consider how you view your future, and how you feel about yourself. Consider to what extent you may feel self-critical, whether you are plagued with guilty feelings, and whether you have been worrying more than usual – for example, about your finances or your health. |
| A |  | **11. Health concerns.** Here we want to know if you worry about your health, regardless of whether you suffer from any known disease or not. |
| A, C | W | **12. Worry about minor things.** The question concerns the extent to which you worry about minor things, worry in advance, or are overly anxious. Consider especially how intense your anxiety is, how often you feel it, and how much effort it takes to disregard it. Note that if you feel worry in certain situations, which you therefore try to avoid, don’t mark that anxiety here but in the next question, Phobias. |
| A | SF | **13. Phobias.** With phobias we mean an exaggerated fear of certain situations that one therefore tries to avoid, not because the situation would be dangerous, but to avoid being scared or embarrassed. Consider if you have any such phobias. It could be taking the underground or bus, being in crowds (shopping malls, queuing, in the cinema), being trapped (e.g., in an elevator) or simply being alone. It can also be feeling uncomfortable in the company of others, at meals with and in similar situations. Also consider how you act, and what you do to avoid the situation. |
| C | C | **14. Obsessions.** Obsessions are recurring, distressing, or frightening thoughts or doubts that intrude even though you don’t want them to, and even though you rationally know they are wrong, unreasonable, or unhealthy. We want to know if you have any such thoughts, how distressing you find them, and how much they affect your everyday life. |
| C | C | **15. Compulsions.** By compulsions we mean things that one feels one must do even though they are unreasonable or exaggerated, and even though one does not want to, to avoid an inner feeling of growing discomfort. Compulsions can be rational, but one performs them when they are not really necessary, or repeats them many times even though once should be enough. We want to know if you are plagued by such compulsions, and how much they impact your everyday life. |
| A | SF | **16. Bodily discomfort.** The question concerns bodily discomfort that can arise when one is anxious, such as heart palpitations, sweating, trouble breathing normally, dizziness, feeling unsteady, cold hands or feet, dry mouth, upset stomach, gas, diarrhoea, increased need to urinate, and so on. Such symptoms can of course occur without feeling very anxious, and also in relation to different diseases. We want to know how distressing these symptoms are, regardless of their cause. |
| A | SF | **17. Aches and pains.** The question concerns if, and how much, you suffer from aches or pain in your body. How often are you in pain, and how intense does it get? Do you need to take pain killers? When you answer the question, you don’t need to take into consideration if you suffer from any known disease or not. Regardless of the causes of the aches and pain, mark the description below that best fits your experience. |
| A | SF | **18. Panic attacks.** By panic attacks, we mean a sudden feeling of strong bodily discomfort, combined with intense fear, usually alongside a feeling of being about to faint, have a heart attack, or losing one’s mind. The attack develops very quickly, in less than a minute, and recedes more slowly. During the attack, one can have heart palpitations, get dizzy, get pins and needles in hands or feet, have trouble breathing, or other bodily symptoms. Think about how distressing any such attacks have been and if you have felt an immediate need to get help (company, medicine). Note that anxiety and worry that does not occur in clear attacks should be marked under question 2, Anxious feelings, not here. |
| D | D | **19. Suicidal thoughts.** This item concerns your appetite for life, and whether you have felt listless and weary of life. Have you had thoughts of suicide, and if so to what extent do you consider it a realistic escape? |

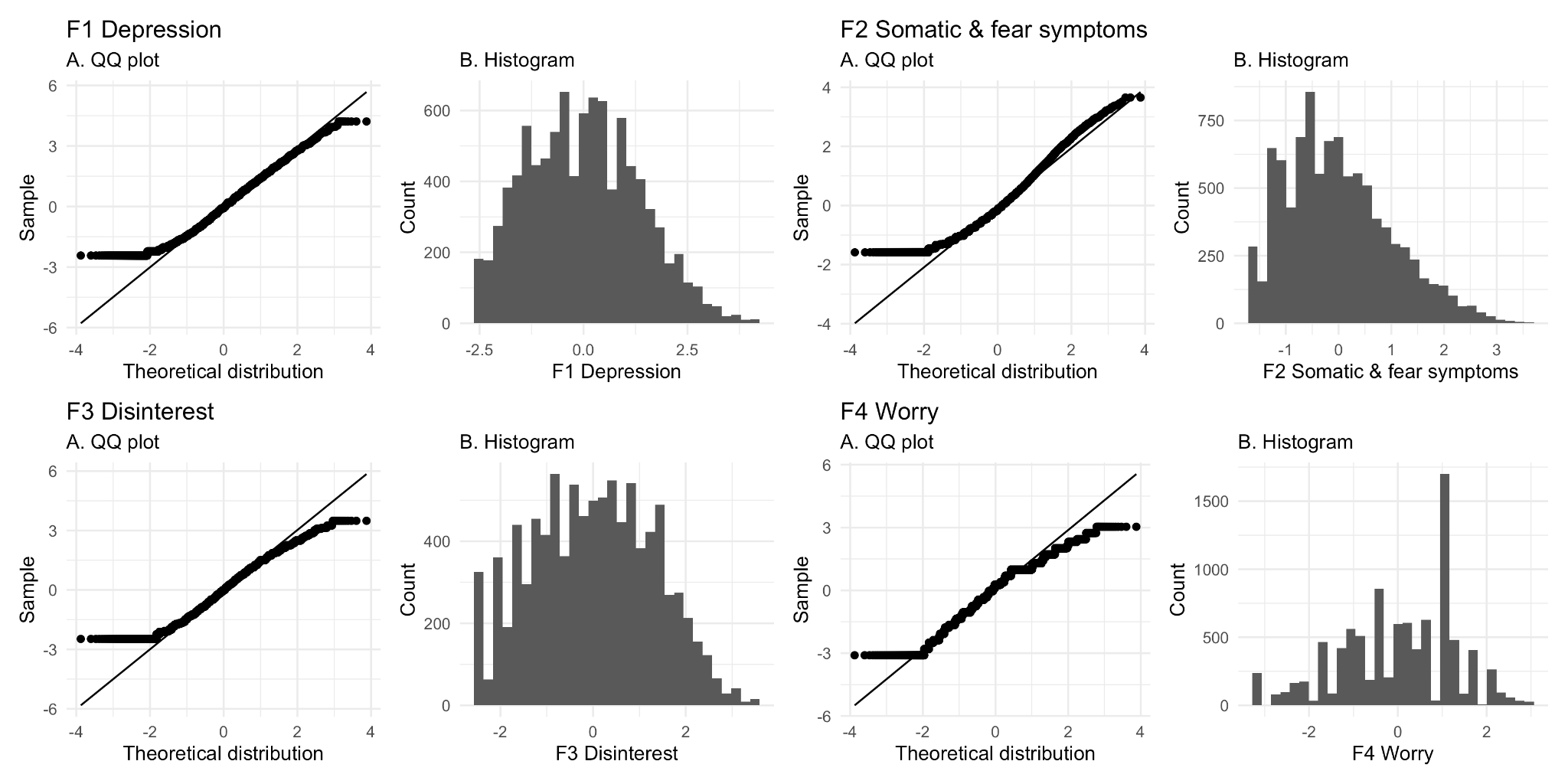

**Supplementary Figure S1. Distribution of the factor scores**

QQ plots and histograms of the four factor scores.
